# Medical Evaluation of Unanticipated Monogenic Disease Risks Identified through Newborn Genomic Screening: Findings from the BabySeq Project

**DOI:** 10.1101/2022.03.18.22272284

**Authors:** Robert C. Green, Nidhi Shah, Casie A. Genetti, Timothy Yu, Bethany Zettler, Talia S. Schwartz, Melissa K. Uveges, Ozge Ceyhan-Birsoy, Matthew S. Lebo, Stacey Pereira, Pankaj B. Agrawal, Richard B. Parad, Amy L. McGuire, Kurt Christensen, Heidi L. Rehm, Ingrid A. Holm, Alan H. Beggs, the BabySeq Project

## Abstract

Genomic sequencing of healthy newborns to screen for medically important genetic information has long been anticipated but data around downstream medical consequences are lacking. Among 159 infants randomized to the sequencing arm in the BabySeq Project, an unanticipated monogenic disease risk (uMDR) was discovered in 18 (11.3%). We assessed uMDR actionability by visualizing scores from a modified ClinGen Actionability SemiQuantitative Metric and tracked medical outcomes in these infants for 3-5 years. All uMDRs scored as highly actionable (mean 9, range: 7-11 on a 0-12 scale) and had readily available clinical interventions. In 4 cases, uMDRs revealed unsuspected genetic etiologies for existing phenotypes, and in the remaining 14 cases provided risk stratification for future surveillance. In 8 cases, uMDRs prompted screening for multiple at-risk family members. These results suggest that actionable uMDRs are more common than previously thought and support ongoing efforts to evaluate population-based newborn genomic screening.

---

Recent advances in the clinical deployment of genome-scale sequencing now make it possible to determine an infant’s complete genome sequence shortly after birth, enabling the identification of deleterious variants associated with monogenic disorders along with other health-related traits^1^. Genomic sequencing as a screening tool in newborns is being explored^2^ and supplementary genomic screening panels are now offered by several commercial laboratories^3^, but there are a number of evidentiary, ethical, and cost concerns that must be considered^1,4-7^. The BabySeq Project is a series of randomized clinical trials of newborn screening by genomic sequencing (GS) that has generated empirical data on mechanisms of consent, gene curation, variant interpretation, disclosure methods, as well as medical, behavioral and economic outcomes, with institutional review board approval and informed consent from all participants^8-13^. In the initial pilot of the project, we recruited infants from both a well-baby nursery and intensive care units (ICU) who were randomized to receive either standard of care newborn screening (NBS) or NBS plus GS. Participants randomized to GS underwent whole exome sequencing (WES) with clinical reporting of pathogenic or likely pathogenic variants (PLPVs) for any genetic condition that was childhood-onset and highly penetrant, or childhood-actionable and at least moderately penetrant^9,14^. A limited subset of actionable adult-onset only conditions were also offered to be returned to families^12^. Results were disclosed to participants’ parents in a counseling session and a disclosure letter was delivered to the family and the newborn’s clinician(s). After disclosure, the study team monitored utility of, and reactions to, the genomic findings by chart review and parental and physician surveys. Parental saliva samples were also collected at enrollment and variant specific testing was completed and disclosed for the parents of all infants with an identified uMDR. The randomized trial was powered to compare psychosocial outcomes and primary results were previously reported.^10^ In this report, we examine the subset of newborns who had findings associated with an unanticipated monogenic disease risk (uMDR) and describe the specific genes and variants detected and disclosed, assess their degree of actionability, and document subsequent medical evaluations and outcomes for these at-risk infants and families.

Of the 325 newborns enrolled in the pilot phase of the BabySeq Project,159 underwent WES and of these, 18 (11.3%) were found to have a PLPV in a gene associated with a childhood-onset-actionable condition, or an actionable adult-onset condition (Table 1)^14^. uMDRs were discovered in 11 of 127 (8.7%) newborns recruited from the well-baby nursery, and 7 of 32 (21.8%) newborns recruited from the ICU. uMDRs in the ICU were not associated with the presenting symptoms and signs of the infants.

**Table 1:**
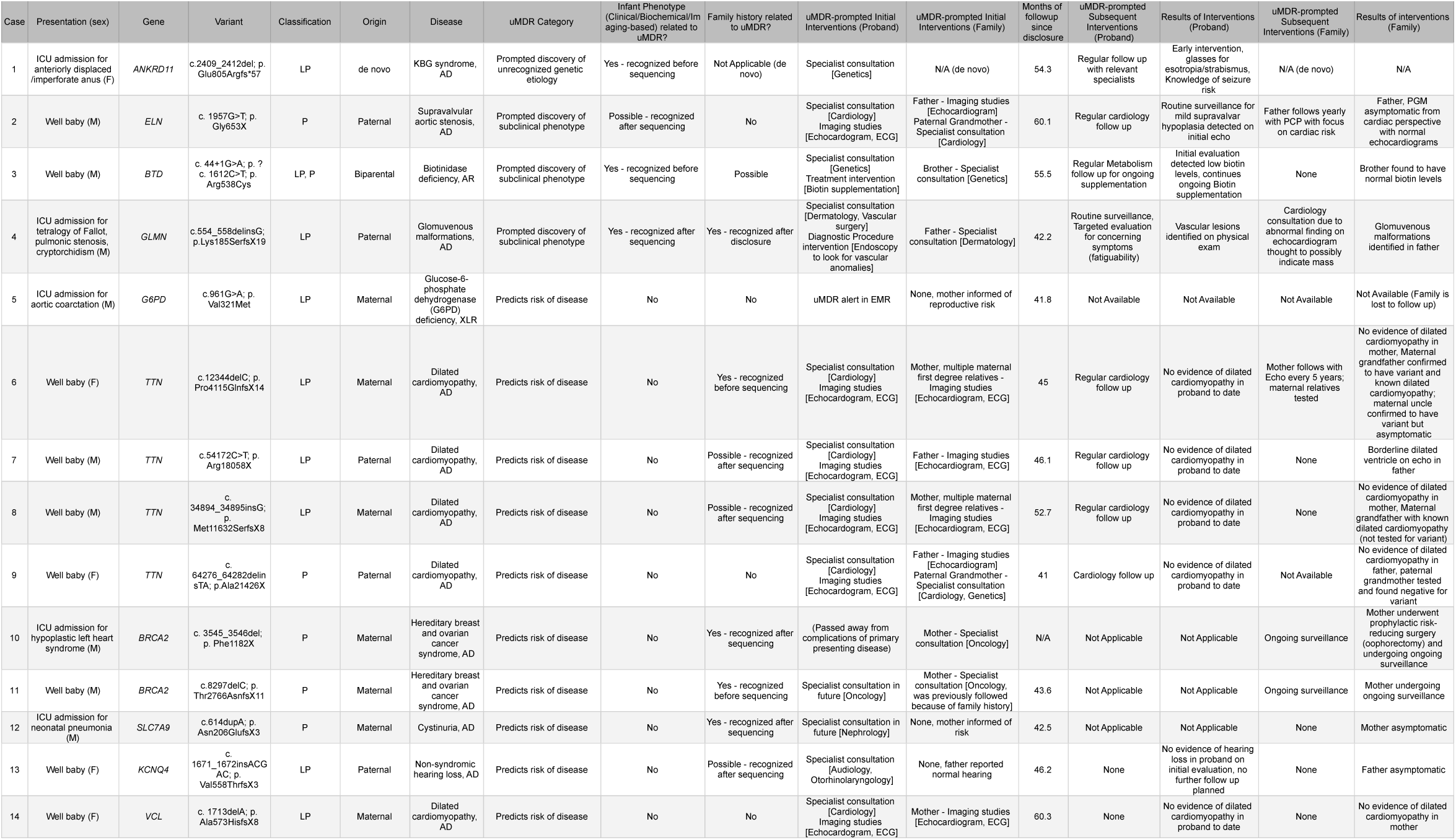
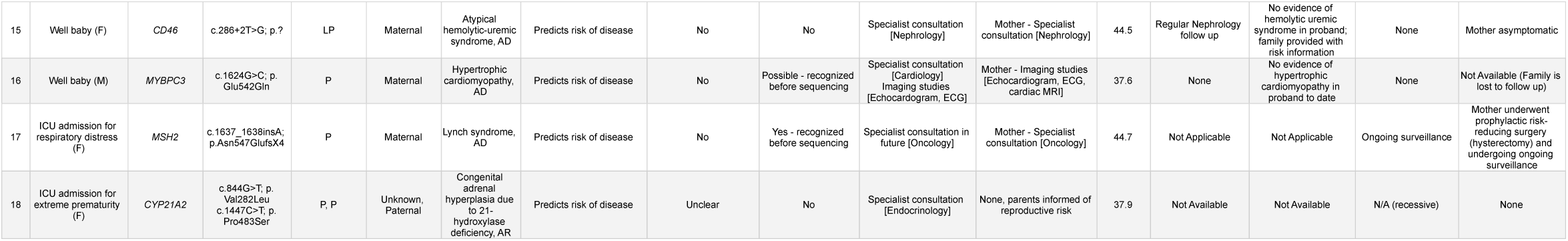
Demographic and clinical characteristics (including phenotype and family history information) of 18 infants witn unanticipated monogenic disease risk (uMDRs), alonf with medical outcomes in probands as well as family members. AD/AR/XLR = autosomal dominant/autosomal recessive/X-linked recessive; P/LP = pathogenic/likely pathogenic varients, hx = history, dx = diagnosis, EMR = electronic medical record

Among the 18 infants with uMDRs, we identified PLPVs in 14 unique genes. We detected heterozygous variants in 11 genes associated with autosomal dominant inheritance; a hemizygous variant in 1 gene (*G6PD*) associated with X-linked recessive inheritance, and compound heterozygous variants in two genes (*BTD* and *CYP21A2*) associated with autosomal recessive inheritance. One *de novo* variant (in *ANKRD11*) provided an unsuspected molecular explanation for a syndromic phenotype under active evaluation. All other variants were inherited. In 3 infants, genomic findings prompted discovery of a clinical phenotype that had not been previously recognized (*ELN, BTD*, and *GLMN*, Table 1), for example echocardiogram revealed mild but abnormal aortic stenosis in the infant with a PV in *ELN* (case 2). In the remainder of the cases (14/18) molecular findings were associated with future disease risk with PLPVs in the following genes: *TTN* in 4 cases; *BRCA2* in 2 cases; and *G6PD, SLC7A9, KCNQ4, CD46, VCL, MYBPC3, MSH2*, and *CYP21A2* with 1 case each. In 8 of those 14 cases, post-disclosure review of the newborn’s family history raised the possibility of additional previously unsuspected and undiagnosed family members with the condition. For example, the maternal grandfather of an infant with a maternally inherited likely pathogenic variant in *TTN* (case 6) had previously been clinically diagnosed with dilated cardiomyopathy and, as a result of the BabySeq finding, was confirmed to carry the same variant. Consequently, the infant’s mother is now routinely followed by echocardiography.

Clinical actionability of uMDRs was classified using the SemiQuantitative Metric (CASQM) adapted in modified form from the ClinGen Actionability Working Group (Supplemental Table 1)^15,16^. The CASQM provides a score of 0-3 on four domains: disease severity, likelihood of developing the disease, efficacy of intervention, and nature of intervention when the patient is carrying the deleterious variant^15^. We scored each of our findings on these top-level concepts, modified the labels for clarity and visualized each of the uMDRs on a radar graph for easier visual interpretation as shown in Figure 1. We also separately assessed the available knowledge base regarding all domains with a score between 0-3^17^. The uMDRs discovered in our study had a mean score of 9, out of a maximum possible score of 12 (range: 7-11). Clinical interventions are available for all the 14 conditions found in the 18 infants, ranging from the initiation of surveillance for cancer risk, hearing loss, and cardiac abnormalities, to biotin supplementation in the case of partial biotinidase deficiency.

**Figure 1:**
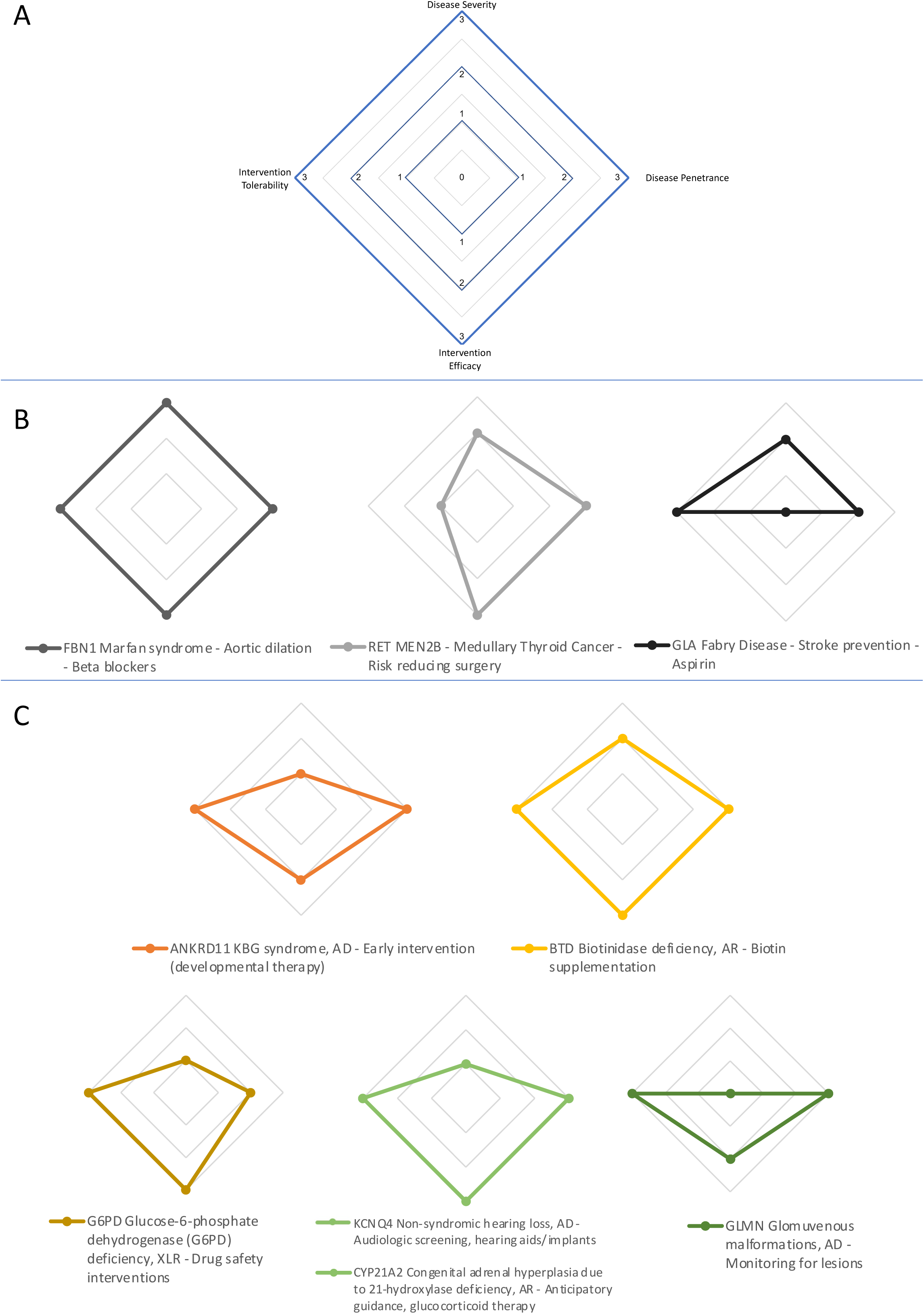
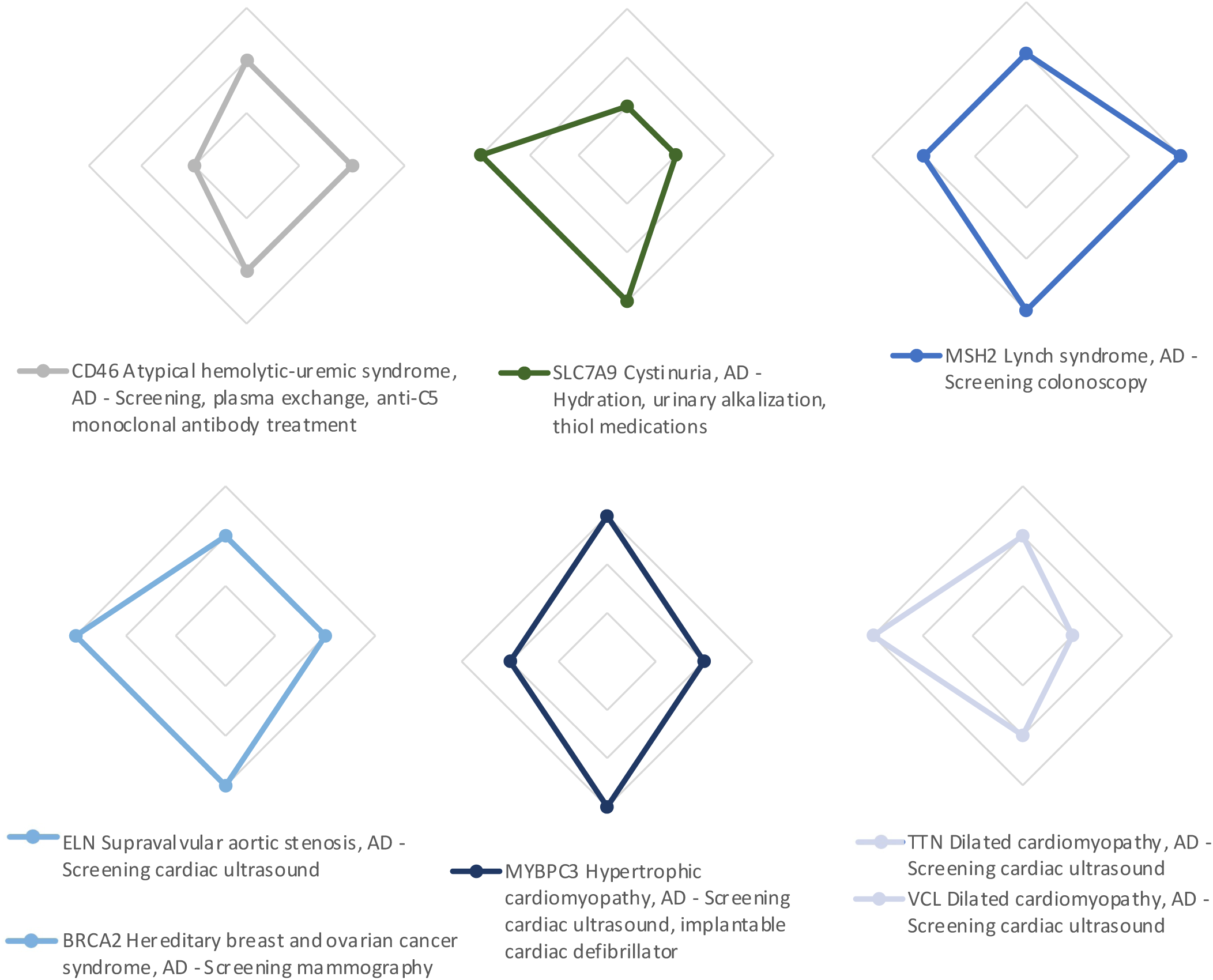
Radar plots that illustrate the pattern of clinical actionability in the specfic genes in which unanticipated pathogenic and likely pathogenic variants (PLPVs) were identified. The plot utilizes a modified Semi-Quantitative Metric adapted from ClinGen as described in the text and online methods. As shown in Panel A, the four points of the diamond on the radar graph (starting from the top and moving clockwise with each figure) represent severity of the fully expressed genetic condition, the penetrance or likelihood that the condition will manifest over an individual’s lifetime, the effectiveness of the specific intervention shown in the figure and the tolerability of the intervention, i.e, its burden and acceptability to patients. A radar plot with maximum area within the diamond would represent a severe genetic condition that has high penetrance with a highly effective intervention that is particularly acceptable to patients because it is minimally invasive or dangerous. Panel B shows sample clinical actionability radar plots for three genes from the ACMG secondary findings list. Panel C demonstrates clinical actionability of the 14 genes in which PLPVs were identified in the 18 infants with an unanticipated monogenic disease risk. Colors indicate different genes.

The infants with uMDRs, and their families, were followed for a median of 44.7 months after disclosure (range: 37.6 to 60.3 months). We reviewed medical records to ascertain what actions were taken for these participants based on their genomic findings (Table 1). We also conducted semi-structured phone interviews with parents 3 years after the conclusion of the study to elicit any relevant information that was not captured in the medical record. We tracked all medical interventions taken on behalf of the infant, as well as medical interventions taken by other family members as a result of the uMDR findings in the sequenced infant. In all but one case (*G6PD* deficiency), the disclosure of uMDR findings generated recommendations for specialist consultation for the child and/or at least one of the parents. Two-thirds (12/18) of the infants received immediate referrals for specialist care, of which 11 completed an initial referral with 9 of the 11 continuing to receive ongoing specialty care. In one-sixth of these (3/18) cases, an adult-onset uMDR was identified in the infant, and parents were encouraged to alert their child when older to seek appropriate screening and care in adulthood. Confirmatory parental testing was performed for all infants with a uMDR, and in 72% of the cases (13/18), genomic findings prompted specialist evaluations and/or diagnostic procedures for one or more family members. To date, at least 4 of these family member evaluations (Cases 4, 6, 7 and 8 in Table 1) have led to medically significant findings. Of the 6 parents discovered to have genetic cardiovascular risk through their infant’s uMDR findings, all 6 had screening workups, and 2 continue to receive ongoing surveillance and care. Of the 3 parents discovered to have genetic cancer risk through their infant’s uMDR findings, all 3 are undergoing ongoing enhanced surveillance, and 2 of the 3 opted for risk-reducing surgeries.

Our study found that 18 out of 159 (11.3%) infants sequenced had a uMDR for a childhood-onset or adult-onset condition, all of which were actionable on a semi-quantitative metric of clinical actionability. In 4 of these cases, the genotype led to the discovery of a phenotype that had not been considered genetic or had not been previously noted, and in 8 of these cases, the genotype findings led to discovery of hitherto unknown at-risk family members. In the 37-60 months following disclosure of the unanticipated genetic findings, two-thirds of the infants and all of their at-risk first-degree family members with uMDRs were referred for specialty consultation, surveillance, or treatment.

Among the 18 infants identified with uMDRs, 3 were subsequently found to have previously unrecognized symptoms or laboratory measures consistent with the underlying condition that had not been previously recognized, and thus were penetrant - albeit with mild or subclinical features. This finding highlights how difficult it is to determine the true penetrance of most monogenic conditions and the importance of considering differences in expressivity over time. While it is well recognized that estimations of penetrance for many genetic conditions are biased toward higher estimates because clinicians are more likely to recognize genetic disease in families with robust inheritance patterns, our findings suggest that penetrance may also be grossly underestimated when genetic diseases present with milder or subclinical symptoms, or in families without obvious family history. Indeed, the very concept of penetrance depends on which particular phenotype is being measured. For example, in an epidemiological study where subclinical features include asymptomatic thickening of the cardiac septum or electrophysiological abnormalities, penetrance around the early features of cardiomyopathy could be recognized earlier than in a study where the first signs of the disease were exercise intolerance or sudden cardiac death^18^.

In the BabySeq Project, we evaluated variants in all disease-associated genes in the genome but only reported variants from those in genes that met our criteria, including 954 pre-curated genes selected for definitive and strong disease-gene association and high penetrance regardless of actionability (884) or moderate evidence or moderate penetrance but high actionability (70)^9^. In addition, 7 genes for adult-onset hereditary breast and ovarian cancer (HBOC) and colon cancer syndromes were offered to all parents. Since the majority of genes listed were selected without regard for actionability, it was surprising to find that all the uMDR discovered in sequenced infants were associated with conditions that were scored as actionable on a modified version of the CASQM. This suggests that the differential in results disclosed between “parents who just want actionable information” and “parents who want any information that is of medical significance to their infant” may not be as large as previously considered.

The concept of actionability is difficult to discuss with patients or research participants, and sometimes even with clinicians and researchers, because for some, the ability to provide enhanced surveillance or even knowledgeable anticipation are considered actionable, while to others this concept only includes conditions where a treatment can slow or stop progression, or unequivocally improve the patient’s prognosis. Figure 1 illustrates how the modified CASQM domains can be represented visually so as to facilitate interpretation. A full diamond with score of 3 in each domain represents the most actionable condition whereas alternative shapes can signal variations in which, for example, penetrance is expected to be lower, or treatment is expected to be more burdensome. This type of visualization could eventually be adapted in a way which would allow clinicians and patients to intuitively understand the specific pros and cons around the actionability of the genetic information they are choosing or are receiving.

Two-thirds of the cases identified with uMDRs were seen by specialists knowledgeable about the genetic condition, and 72% of the cases prompted specialist evaluations and/or follow-up procedures for one or more family members. This speaks to the benefits of alerting family members to a genotype carried by one individual and to the powerful advantages of cascade testing, a phenomenon that has been recognized as far more efficient and cost-effective than population screening^19,20^, but that is implemented inconsistently and with variable success in clinical genetics today^21^.

The BabySeq Project is the first rigorous examination of uMDR findings in an unselected cohort of newborns and has several limitations. Participation in our project was offered to the parents of several thousand infants born in a tertiary care medical center, and those who enrolled were more educated and of higher socio-economic status than the general population^10,13^. The sample size of infants carrying uMDRs is small and the constellation of unanticipated genetic findings cannot be presumed to be representative of what might be found in population-level sequencing. In 5 cases of uMDR disclosure, neither the physical examination, subsequent testing, nor the family history revealed any new or useful information, leaving these 5 families with risk information that may be less likely to lead to penetrant disease. A much larger study with a more diverse population would be needed to ascertain whether the anxiety to parents and cost to the healthcare system associated with specialty consultations and surveillance around these risks is worthwhile. However, it is reassuring that in a separate report we recently demonstrated that neither sequencing, nor receiving uMDRs, was associated with significantly greater parental distress or disruption of the parent-child bond, and that parents felt empowered by both positive and negative results^10^. In assessing the reactions of parents in a study like this, it is important to keep in mind that parents self-select to enroll, and their self-selection reflects a desire to receive risk information and may be biased toward greater optimism around the results.

In summary, these early medical outcomes from the BabySeq Project suggest that a large proportion of infants identified as having unanticipated monogenic disease risk had actionable conditions and in nearly all cases the genomic findings resulted in a change in medical care.

## Data Availability

All data produced in the present study are available upon reasonable request to the authors.

## ONLINE METHODS

The BabySeq Project is a series of randomized controlled trials of whole exome sequencing (WES) in newborns from the newborn nursery (NBN) or intensive care unit (ICU) who were randomized to conventional care plus family history assessment and genetic counseling alone or with WES. Detailed methodology for the study design and recruitment can be found elsewhere^1,2^. In brief, parents and newborns from the well-baby nursery at Brigham and Women’s Hospital (BWH) and parents and sick newborns from the neonatal intensive care unit and other intensive care units (NICU/ICUs) at BWH, Boston Children’s Hospital (BCH), and Massachusetts General Hospital (MGH), were approached, consented, and enrolled into the study regardless of the absence or presence of a specific phenotype. A blood sample was obtained from each newborn for DNA isolation and analysis whereas parent(/s) provided saliva samples. Within each cohort, healthy and sick, the families were randomized to receive family history and standard newborn screening (NBS) [the control arm] or to the modified standard of care plus exome sequencing (ES) [the ES arm]. Both arms had a three-generation family history collected and evaluated by a study genetic counselor. Additionally, parental surveys at enrollment (baseline), immediately post-disclosure, and 3 and 10 months post-disclosure were administered.

### Gene Selection and Variant Analysis

Detailed description of the rationale for gene selection and variant analysis in the BabySeq Project have been previously published^3,4^. In brief, exomes were annotated, filtered and results analyzed to identify pathogenic or likely pathogenic variants (PLPVs) that met criteria for return, including variants found in genes with definitive or strong disease-gene association and high penetrance regardless of actionability or variants found in genes with moderate evidence or moderate penetrance but high actionability. Whole-exome sequencing (WES) was performed at the CLIA/CAP-accredited Clinical Research Sequencing Platform of the Broad Institute of MIT and Harvard, and Sanger confirmation was performed at the CLIA-accredited Mass General Brigham Laboratory for Molecular Medicine as previously described. All nGS results were returned to parents and placed into the medical record via a Newborn Genomic Sequencing Report (NGSR), which included an indication-based analysis (IBA) for any additional diagnostic assessment related to a clinical indication.

### Disclosure protocol

Parents attended a disclosure session facilitated by a study physician and genetic counselor, on average four months after enrollment (range: 1.2 to 10.2 months), where they were informed of their randomization status and given their family history report. Parents in the ES arm also received the NGSR, and results of an IBA (if performed). At disclosure, the study physicians (who are all trained in either clinical genetics, neurology and/or neonatology) performed detailed physical examination to identify features that might have been previously missed. After disclosure of results, the genetic counselor and physician prepared a note summarizing the visit. These notes, along with the family history report, NBS report, and, for those in the sequencing arm, the NGSR/IBA, were then mailed to the parents and faxed to the infant’s pediatrician and other providers and all these documents were uploaded to the infant’s medical record at BWH or BCH.

### Health Outcomes in this Analysis

We reviewed all available medical records to ascertain downstream actions taken both for the proband and family members. We also conducted phone interviews 46.8 months (mean; range: 37.6 to 60.3 months) after the conclusion of the study to identify any relevant information that may not have been captured in the medical records. We collected a range of outcomes to study the process and effect of integrating WGS into primary care. We evaluated downstream healthcare use attributable to the genomic results both for primary care as well as specialty care. We also evaluated family member cascade testing and family member health care utilization (where available).

### Actionability Analysis

Clinical severity of potential conditions identified and available interventions were graded by modifying the ClinGen Actionability Semiquantitative Metric (SQM)^5^. In the BabySeq Project, the SQM was applied to assess potential actionability over the lifetime of a newborn screened as a primary quest for identifying childhood-onset or childhood-actionable condition, or an actionable adult-onset condition, all of which have clinical interventions available. Hence the ClinGen Actionability SQM was adapted such that for each gene-disease pair, we scored outcome-intervention pairs for outcomes with relatively high penetrance when more than one outcome was possible. Additionally, we scored outcome-intervention pairs based on all available evidence as opposed to only pediatric actionability, given the important cascade testing implications.

## Data Availability

The data/analyses reported in this paper have been deposited in the Newborn Screening Translational Research Network Longitudinal Pediatric Data Resource under accession identifier nbs000002.v1.p1.

## Code Availability

Not applicable

## Author Disclosures

Dr. Green has received compensation for advising Genomic Life and is co-founder of Genome Medical.

Dr. Shah is a member of the Scientific Advisory Board for Neuberg Center for Genomic Medicine.

Ms. Genetti has received compensation for consulting for Kate Therapeutics.

Dr. Yu has consulted and received compensation or honoraria from Eisai, BioMarin, GeneDx, Takeda, and Alnylam.

Ms. Zettler has received compensation for consulting for Novartis Gene Therapies.

Dr. Lebo is employed by a not-for-profit, fee-for-service clinical laboratory at Mass General Brigham offering genomic screening.

Dr. Agrawal is a member of the Scientific Advisory Board for GeneDx and Illumina, Inc.

Dr. McGuire is a member of the board of the Greenwall Foundation and is on the Scientific Advisory Board for Danaher Life Sciences, Geisinger Research, and the Morgridge Institute for Research.

Dr. Rehm is a member of the Scientific Advisory Board for Genome Medical and is employed as the medical and clinical laboratory director for a fee-for service laboratory offering genomic sequencing at the Broad Institute of MIT and Harvard.

Dr. Holm is a member of the Scientific Advisory Board for Biomarin for vosoritide.

Dr. Beggs has received funding from the NIH, MDA (USA), AFM Telethon, Alexion Pharmaceuticals Inc., Audentes Therapeutics Inc., Avidity Biosciences, Dynacure SAS, and Pfizer Inc. He has consulted and received compensation or honoraria from Asklepios BioPharmaceutical Inc, Audentes Therapeutics, Biogen, F. Hoffman-La Roche AG, GLG Inc, Guidepoint Global, and Kate Therapeutics, and holds equity in Kinea Biosciences and Kate Therapeutics.

**Supplementary Table 1.** Clinical actionability of specific genes in which pathogenic and likely pathogenic variants (PLPVs) were identified in infants with uMDR. The table illustrates the score assignment based on the modified semi-Quantitative Metric adapted from ClinGen as described in the text and online methods. Where available, the ClinGen Actionability Scores as well as age-related semi-quantitative metric actionability scores from the NC NEXUS study (with NC NEXUS knowledge base scores in brackets) are also shown to facilitate comparison

| Supplementary Table 1. |  |  |  |  |  |  |  |  |  |  |  |
| --- | --- | --- | --- | --- | --- | --- | --- | --- | --- | --- | --- |
| Case | Presentation | Gene | Disease-Intervention | Severity | Likelihood of disease | Efficacy of intervention | Burden of intervention | Total score | Knowledge Base Strength | Clingen Actionability Score (where available) | NC NEXUS ASQM Score (ASQM knowledge base score) |
| 1 | ICU admission for anteriorly displaced /imperforate anus | ANKRD11 | KBG syndrome, AD - Early intervention (developmental therapy) | 1 | 3 | 2 | 3 | 9 | 1 | - | - |
| 2 | Well baby | ELN | Supravalvular aortic stenosis, AD - Screening cardiac ultrasound | 2 | 2 | 3 | 3 | 10 | 3 | - | 11 (2) |
| 3 | Well baby | BTB | Biotinidase deficiency, AR - Biotin supplementation | 2 | 3 | 3 | 3 | 11 | 3 | 11CC | 10 (3) |
| 4 | ICU admission for tetralogy of Fallot, pulmonic stenosis, cryptorchidism | GLMN | Glomovenous malformations, AD - Monitoring for lesions | 0 | 3 | 2 | 3 | 8 | 1 | - | - |
| 5 | ICU admission for aortic coarctation | G6PD | Glucose-6-phosphate dehydrogenase (G6PD) deficiency, XLR - Drug safety interventions | 0 | 3 | 3 | 3 | 9 | 3 | - | 9 (3) |
| 6 | Well baby | TTN | Dilated cardiomyopathy, AD - Screening cardiac ultrasound | 2 | 1 | 2 | 3 | 8 | 2 | - | 9 (2) |
| 7 | Well baby | TTN | Dilated cardiomyopathy, AD - Screening cardiac ultrasound | 2 | 1 | 2 | 3 | 8 | 2 | - | 9 (2) |
| 8 | Well baby | TTN | Dilated cardiomyopathy, AD - Screening cardiac ultrasound | 2 | 1 | 2 | 3 | 8 | 2 | - | 9 (2) |
| 9 | Well baby | TTN | Dilated cardiomyopathy, AD - Screening cardiac ultrasound | 2 | 1 | 2 | 3 | 8 | 2 | - | 9 (2) |
| 10 | ICU admission for hypoplastic left heart syndrome | BRCA2 | Hereditary breast and ovarian cancer syndrome, AD - Screening mammography | 2 | 2 | 3 | 3 | 10 | 3 | 8-10AA | 10 (3) |
| 11 | Well baby | BRCA2 | Hereditary breast and ovarian cancer syndrome, AD - Screening mammography | 2 | 2 | 3 | 3 | 10 | 3 | 8-10AA | 10 (3) |
| 12 | ICU admission for neonatal pneumonia | SLC7A9 | Cystinuria, AD - Hydration, urinary alkalization, thiol medications | 1 | 1 | 3 | 3 | 8 | 2 | - | 8 (2) |
| 13 | Well baby | KCNQ4 | Non-syndromic hearing loss, AD - Audiologic screening, hearing aids/implants | 1 | 3 | 3 | 3 | 10 | 2 | - | 10 (2) |
| 14 | Well baby | VCL | Dilated cardiomyopathy, AD - Screening cardiac ultrasound | 2 | 2 | 2 | 3 | 9 | 2 | - | 10 (1) |
| 15 | Well baby | CD46 | Atypical hemolytic-uremic syndrome, AD - Screening, plasma exchange, anti-C5 monoclonal antibody treatment | 2 | 2 | 2 | 1 | 7 | 2 | - | - |
| 16 | Well baby | MYBPC3 | Hypertrophic cardiomyopathy, AD - Screening cardiac ultrasound | 3 | 2 | 3 | 2 | 10 | 3 | 10NN | 10 (3) |
| 17 | ICU admission for respiratory distress | MSH2 | Lynch syndrome, AD - Screening colonoscopy | 2 | 3 | 3 | 2 | 10 | 3 | 8AA-9AB-10AA | 10 (3) |
| 18 | ICU admission for extreme prematurity | CYP21A2 | Congenital adrenal hyperplasia due to 21-hydroxylase deficiency, AR - Anticipatory guidance, glucocorticoid therapy | 1 | 3 | 3 | 3 | 10 | 3 | - | 11(3) |

